# Modelling the transmission dynamics of COVID-19 in six high burden countries

**DOI:** 10.1101/2020.04.22.20075192

**Authors:** Azizur Rahman, Md Abdul Kuddus

**Affiliations:** Data Science Research Unit, School of Computing and Mathematics, Charles Sturt University, Wagga Wagga, NSW, Australia; Australian Institute of Tropical Health and Medicine, James Cook University, Townsville, QLD, Australia; Department of Mathematics, University of Rajshahi, Rajshahi-6205, Bangladesh

**Keywords:** Epidemic model, nonlinear incidence, transmission rate, COVID-19, simulations

## Abstract

The new coronavirus disease, officially known as COVID-19, originated in China in 2019 and has since spread around the globe. We presented a modified *Susceptible-Latent-Infected-Removed* (SLIR) compartmental model of COVID-19 disease transmission with nonlinear incidence during the epidemic period. We provided the model calibration to estimate parameters with day wise corona virus (COVID-19) data i.e. reported cases by worldometer from the period of 15^th^ February to 30^th^ March, 2020 in six high burden countries including Australia, Italy, Spain, USA, UK and Canada. We estimate transmission rates for each countries and found that the highest transmission rate country in Spain, which may be increase the new cases and deaths in Spain than the other countries. Sensitivity analysis was used to identify the most important parameters through the partial rank correlation coefficient method. We found that the transmission rate of COVID-19 had the largest influence on the prevalence. We also provides the prediction of new cases in COVID-19 until May 18, 2020 using the developed model and recommends, control strategies of COVID-19. The information that we generated from this study would be useful to the decision makers of various organizations across the world including the Ministry of Health in Australia, Italy, Spain, USA, UK and Canada to control COVID-19.

## 1. Introduction

Following the outbreak of novel Severe Acute Respiratory Syndrome Coronavirus-2 (SARS-Cov-2) or COVID-19 constitute a persistent and significant public-health problem across the globe. As of March 30th, 2020, the ongoing global epidemic outbreak of COVID1-19 has spread to at least 180 countries and territories on 6 countries including Australia, Italy, Spain, USA, UK and Canada, resulted approximately 946,876 cases of COVID-19, and 48,137 individuals died from this disease [1]. In the Australia, Italy, Spain, USA, UK and Canada COVID-19 infection and death reached 4460, 101739, 87956, 163788, 22141 and 7448, as well as 30, 11591, 7716, 3143, 1408 and 89 with mortality ratios nearly 0.67%, 11.39%, 8.77%, 1,9%, 6.4% and 1.2% respectively [1]. Figure 1 shows the cumulative number of confirmed cases and deaths of COVID-19 in six selected countries from February 15^th^ to March 30^th^, 2020.

**Figure 1:**
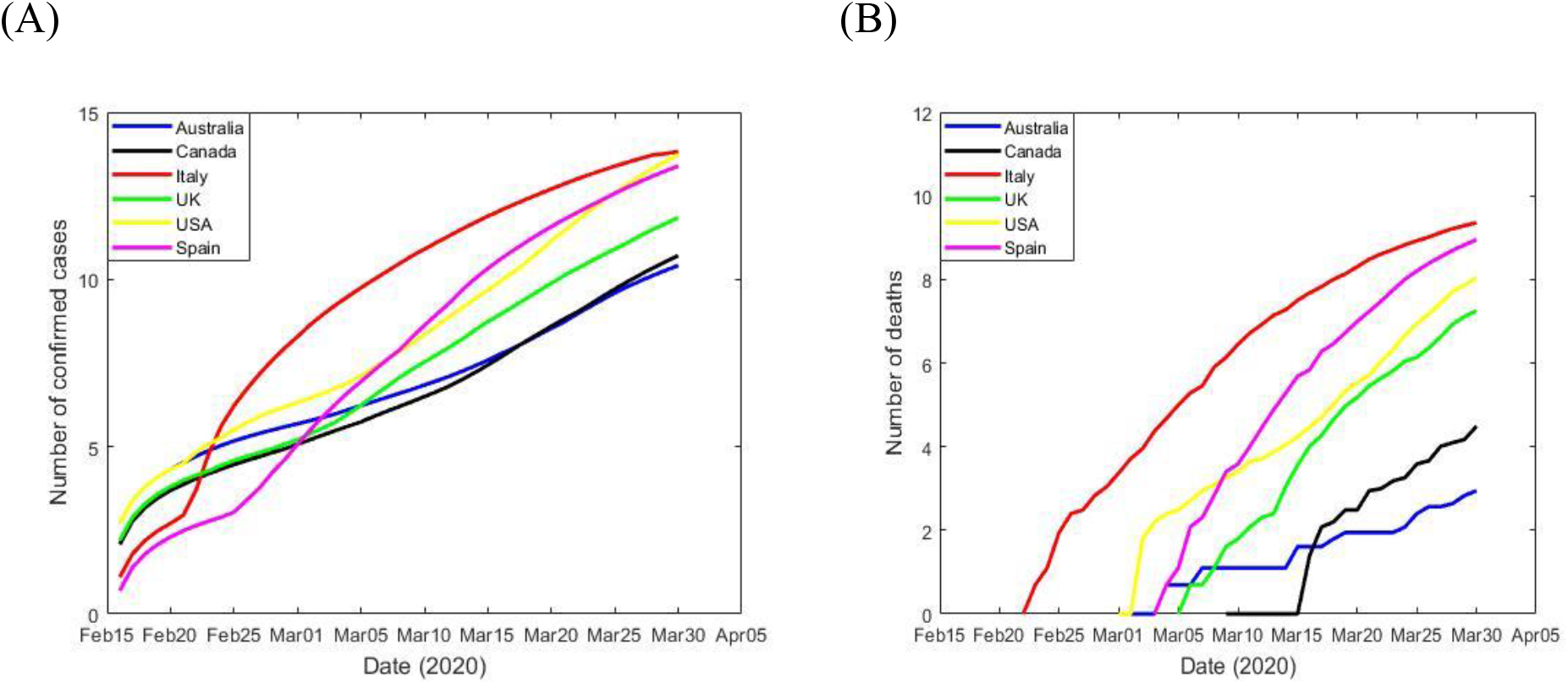
Graphs of six selected countries using a log scale (A)–cumulative number of COVID-19 cases and (B)–cumulative number of COVID-19 deaths.

The highest burden of COVID-19 is not only dependent on the health system but also depend quickly response. For example, in Italy, the first confirmed COVID-19 cases on February 15 and then after few days thousands of people infected by COVID-19. The problem is not that the Italy government didn’t respond to the COVID-19. The problem is that it always responded slightly too slow and with slightly too much moderation. What has resulted in China reveals that quarantine, social distance, and isolation of infected populations can contain the epidemic. This impact of the COVID-19 response in China is advocating for many countries where COVID-19 is starting to spread. However, it is unclear whether other countries can implement the stringent measures China eventually adopted. Singapore and Hong Kong, both of which had severe acute respiratory syndrome (SARS) epidemics in 2002–03, present concern and many lessons to other countries. In both places, COVID-19 has been maintained well to date, notwithstanding early cases, by early government progress and through social distancing patterns used by individuals.

The course of an epidemic is defined by a series of key factors, some of which are poorly understood at present for COVID-19. Mathematical modelling is one of the most powerful tools for infectious disease control that can be used for both predictions about behaviour and for understanding infectious disease dynamics [2-4]. Many researchers have implemented mathematical modelling frameworks to gain insights into different types of infectious diseases [5-9]. Although models can range from very simple to highly complex, one of the commonest practices to improve understanding of infectious disease dynamics is the compartmental mathematical model [10].

In mathematical models, the incidence rate plays an important role in the transmission of infectious diseases. The number of individuals who become infected per unit time is called the incidence rate in the epidemiology perspective [11]. Here, we consider the nonlinear incidence rate due to the number of effective contact between infective and susceptible individuals may saturate at high levels through the crowding of infectives individuals [12]. This model is also used to calibrate and make prediction the number of COVID-19 cases data in six countries including Australia, Italy, Spain, USA, UK and Canada to estimate the model parameters. Sensitivity analysis also performed to identify the most important model parameters that could potentially support policymakers to control COVID-19 outbreak in the selected countries. The model findings can be also helpful to many other countries which are dealing with critical outbreak of COVID-19.

The rest of the paper is structured as follows: Section 2 presents model descriptions. Sections 3 and 4 performed the model calibration and sensitivity analysis. A brief discussion and concluding remarks finalize the paper.

## 2. Model description and analysis

We considered a modified SLIR compartmental model of COVID-19 transmission with nonlinear incidence between the following mutually exclusive compartments: S(t)-susceptible individuals; L(t)-latent individuals who have not yet progressed to active infection; I(t)-infected individuals who are both infected and infectious, and R(t)-recovered individuals who are previously infected but successfully recovered. A typical SLIR model is depicted in Figure 2.

**Figure 2:**
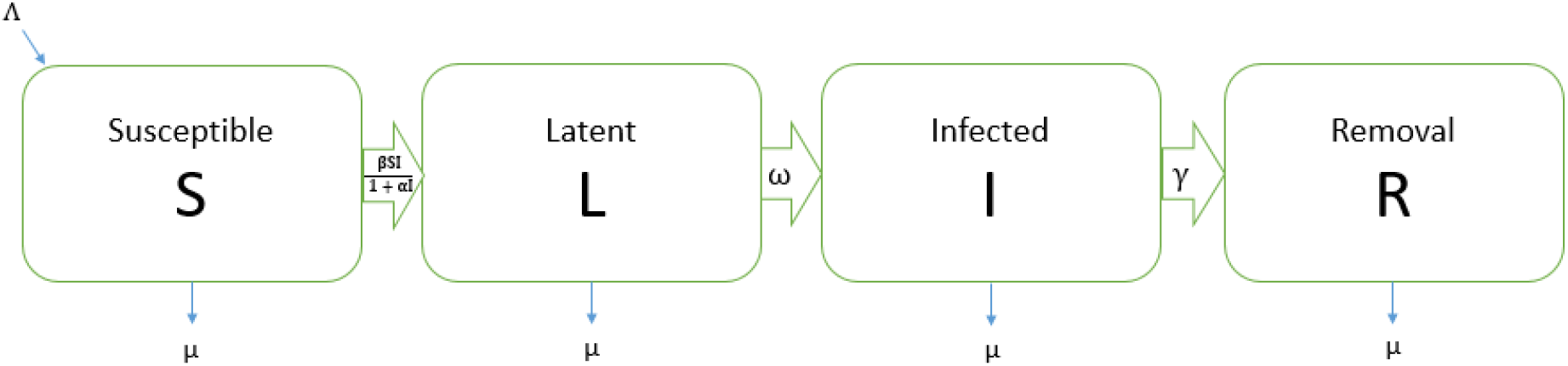
Flow chart of the SLIR mathematical model showing the four states and the transitions in and out of each state. Here, S = Susceptible population, L = Latent population, I = Infected population, R = Removal population, Λ = recruited rate, μ = Death rate, β = Transmission rate, α=Force of saturates infection, ω = Progression rate to active disease, γ = Recovery rate.

Let the susceptible individuals be recruited at a constant rate Λ and they may be infected at a time dependent rate 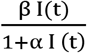. Here, 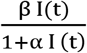 is represents the saturated incidence rate, which tends to a saturated level when I(t) gets large. βI(t) measures the force of infection when the disease is entering a fully susceptible population, and 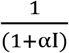 measures the inhibition effect from the behaviour change of susceptible individuals when their number increase or from the effect of risk factors including crowded environment of the infective individuals with α determines the level at which the force of infection saturates. Individuals in the different compartments suffer from natural death at the same constant rate μ. All infected individuals move to the latently infected compartment, L(t). Those with latent infection progress to active infection (the I compartment) as a result of reactivation of the latent infection at rate ω. A proportion of the infected individuals recover through treatment and natural recovery rate γ and move into the recovered compartment R(t). In this case the model can be expressed by the following four differential equations:

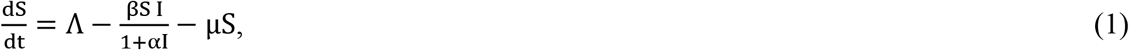

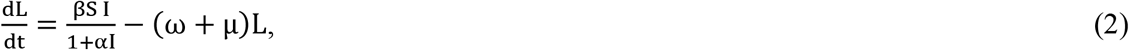

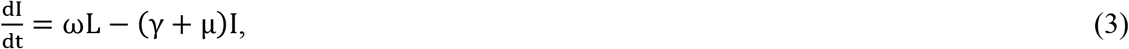

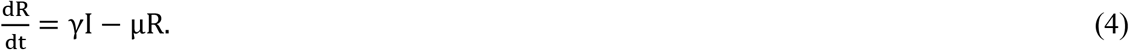

Given non-negative initial conditions for the system above, it is straightforward to show that each of the state variables remain non-negative for all t > 0. Moreover, summing equations (1)-(4) we find that the size of the total population, N(t) satisfies

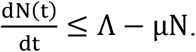

Integrating this equation we find

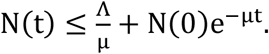

This shows that the total population size N(t) is bounded in this case, and naturally it follows that each of the compartment states (i.e. S, L, and I etc.) are also bounded.

## 3. Estimation of model parameters

In this section we estimated the model parameters based on the available six countries COVID-19 reported cases data from the worldometers.info [1]. Figure 3 presents the curve of cumulative confirmed COVID-19 cases in each day during the period from the 15^th^ February to 30^th^ March 2020 in Australia, Italy, Spain, USA, UK and Canada. In order to parameterise the model (1) – (4), we obtained some of the parameter values from the literature (see, Table 1), other were estimated or fitted from the data. The best-fitted parameter values were obtained by minimizing the error using least-square fitting method between the COVID-19 cases data and the solution of the proposed model (1) – (4) (see, blue solid graph in Figure 3).

**Table 1:** Depiction and estimation of the model parameters for six countries

| Countries | Parameters | Description | Estimated Values | References |
| --- | --- | --- | --- | --- |
| **Australia** | N | Population in 2020 | 254,998,84 | [13] |
| | $\mu$ | Death rate | $\frac{1}{70} \text{ yr}^{-1}$ | [14] |
| | $\beta$ | Transmission rate | $1.3137 \times 10^{-7}$ | Fitted |
| | $\omega$ | Progression rate from L to I | 0.01 | Assumed |
| | $\gamma$ | Recovery rate | 0.1 | Assumed |
| | $\alpha$ | Infection saturates rate | 0.00001 | Assumed |
| | $\Lambda$ | Recruitment rate | 1 | |
| **Italy** | N | Population in 2020 | 604,618,26 | [15] |
| | $\mu$ | Death rate | $\frac{1}{70} \text{ yr}^{-1}$ | [14] |
| | $\beta$ | Transmission rate | $2.5722 \times 10^{-8}$ | Fitted |
| | $\omega$ | Progression rate from L to I | 0.00931 | Assumed |
| | $\gamma$ | Recovery rate | 0.16 | Assumed |
| | $\alpha$ | Infection saturates rate | 0.0000058 | Assumed |
| | $\Lambda$ | Recruitment rate | 1 | |
| **Spain** | N | Population in 2020 | 467,547,78 | [16] |
| | $\mu$ | Death rate | $\frac{1}{70} \text{ yr}^{-1}$ | [14] |
| | $\beta$ | Transmission rate | $6.7616 \times 10^{-7}$ | Fitted |
| | $\omega$ | Progression rate from L to I | 0.00034 | Assumed |
| | $\gamma$ | Recovery rate | 0.3 | Assumed |
| | $\alpha$ | Infection saturates rate | 0.0000004 | Assumed |
| | $\Lambda$ | Recruitment rate | 1 | |
| **USA** | N | Population in 2020 | 331,002,651 | [17] |
| | $\mu$ | Death rate | $\frac{1}{70} \text{ yr}^{-1}$ | [14] |
| | $\beta$ | Transmission rate | $6.1269 \times 10^{-7}$ | Fitted |
| | $\omega$ | Progression rate from L to I | 0.03 | Assumed |
| | $\gamma$ | Recovery rate | 0.028 | Assumed |
| | $\alpha$ | Infection saturates rate | 0.00000266 | Assumed |
| | $\Lambda$ | Recruitment rate | 1 | |
| **UK** | N | Population in 2020 | 678,869,11 | [18] |
| | $\mu$ | Death rate | $\frac{1}{70} \text{ yr}^{-1}$ | [14] |
| | $\beta$ | Transmission rate | $5.9950 \times 10^{-7}$ | Fitted |
| | $\omega$ | Progression rate from L to I | 0.0003 | Assumed |
| | $\gamma$ | Recovery rate | 0.1 | Assumed |
| | $\alpha$ | Infection saturates rate | 0.00001 | Assumed |
| | $\Lambda$ | Recruitment rate | 1 | |
| **Canada** | N | Population in 2020 | 377,421,54 | [19] |
| | $\mu$ | Death rate | $\frac{1}{70} \text{ yr}^{-1}$ | [14] |
| | $\beta$ | Transmission rate | $4.5237 \times 10^{-7}$ | Fitted |
| | $\omega$ | Progression rate from L to I | 0.0003 | Assumed |
| | $\gamma$ | Recovery rate | 0.2 | Assumed |
| | $\alpha$ | Infection saturates rate | 0.00001 | Assumed |
| | $\Lambda$ | Recruitment rate | 1 | |

**Figure 3:**
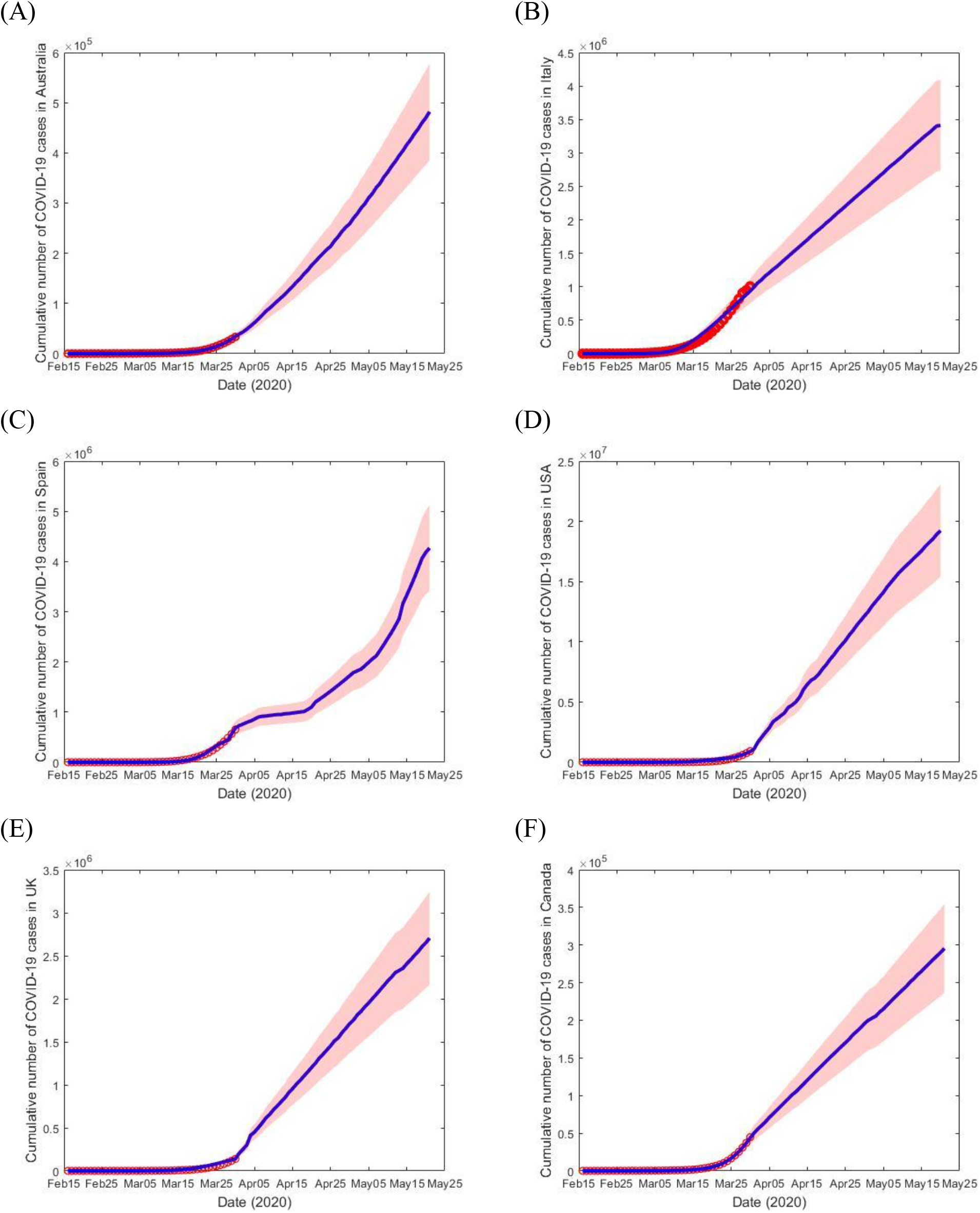
Measured and predicted number of cumulative COVID-19 cases from February 15, 2020 to May 18, 2020 (red dot) in six different high burden countries (A) Australia, (B) Italy, (C) Spain, (D) USA, (E) UK and (F) Canada, and the corresponding model (blue solid curve) with the 95% confidence interval (CI) measure in the blue shaded limits.

The objective function used in the parameter estimation is as follows

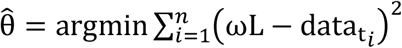

where 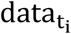 denotes the COVID-19 data and ωL are the corresponding model solution at time t_i_, while n is the number of available actual data points. Findings reveal that the proposed model is well-fitted with the data.

The prediction results from the model are also depicted in Figure 3 to assist in evidenced based decision making process. For example, health department and decision makers including political leaders of Australia, Italy, Spain, USA, UK and Canada those who bear the greatest response for national health systems for what is predicted to happen in the days, weeks and month to come. They can then implement measures regarding staff resources and hospital beds to meet the challenges of this difficult time. However, if the number of infected individuals follows this trend for the next month, there will be more than 400,000 in Australia, 350,000,0 in Italy, 400,000,0 in Spain, 180,000,00 in USA, 270,000,0 in UK and 300,000 in Canada patients infected by May 18, as shown in Figure 3.

## 4. Sensitivity analysis

Sensitivity analysis is performed to investigate the parameters that process the greatest influence on the model outputs [20, 21]. In this study, we performed the partial rank correlation coefficient (PRCC), which is a global sensitivity analysis technique proven to be the most reliable and efficient sampling based method, is utilized [21, 22]. About 100,000 simulations are performed and a uniform distribution is assigned to each model parameter and sampling is performed independently. Positive (negative) correlations suggests that a positive (negative) variation in the parameter will increase (decrease) the model outcome [21]. Here the model outputs we consider are the number of infectious individuals I (where,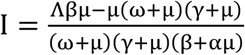) and the basic reproduction number R_0_(where, 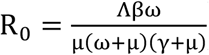). Figure 4 and Figure 5 display the correlation between I and R_0_ corresponding to the model parameters β, ω, α and γ. Parameters β and ω have positive PRCC values, implying that a positive change of these parameters will increases the infectious individuals I and the basic reproduction number R_0_. In contract parameters α and γ have negative PRCC values, which implies that increasing theses parameters will decrease infectious individuals I and the basic reproduction number R_0_.

**Figure 4:**
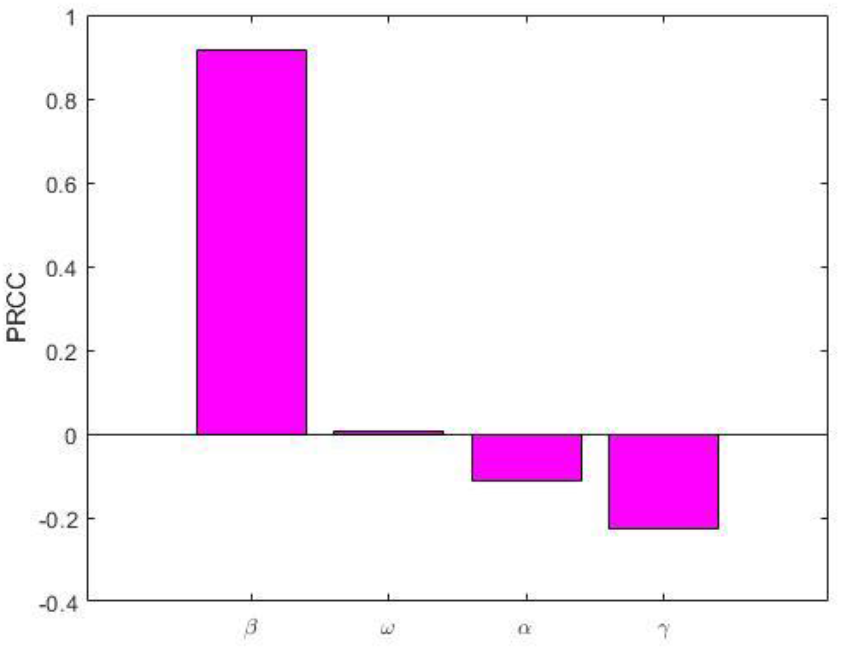
PRCC values depicting the sensitivities of the model output infectious individuals I with respect to the estimated parameters β, ω, α and γ.

**Figure 5:**
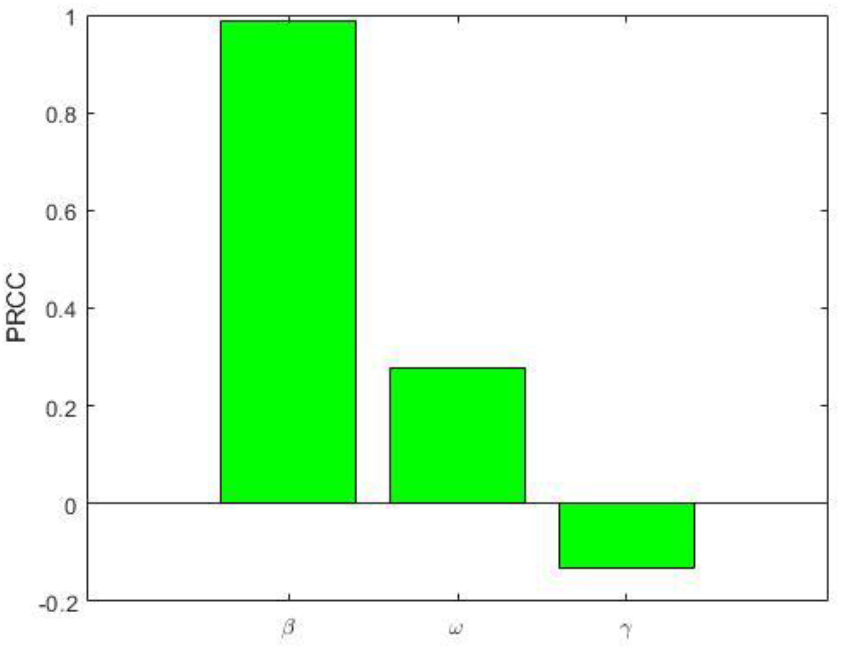
PRCC values depicting the sensitivities of the model output basic reproduction number R_0_ with respect to the estimated parameters β, ω and γ.

## 5. Discussion and Concluding remarks

In this paper, we presented a modified SLIR compartmental model with nonlinear incidence. We estimate number of cases from COVID-19 infection and apply it to data from the COVID-19 epidemics in Australia, Italy, Spain, USA, UK and Canada until 18 May 2020. After model calibration we estimates the transmission rates in Australia, Italy, Spain, USA, UK and Canada are 1.3137 × 10^−7^, 2.5722 × 10^−8^, 6.7616 × 10^−7^, 6.1269 × 10^−7^, 5.9950 × 10^−7^ and 4.5237 × 10^−7^ respectively. The model estimates show a strong relationship with transmission rate and number of cases in COVID-19 of the selected countries.

Within the six different countries we found that Spain has the highest transmission rate than the other selected countries, which may be increase massive number of COVID-19 cases and make worst situation in Spain. We assume that initially Spain government may be not taken proper action to control transmission including handwashing, social distancing and good respiratory hygiene etc. For instance, in China they take immediate action for transmission control including lockdown in every cities that is way they are able to minimize the outbreak of COVID-19. Our finding is consistent with observations, because COVID-19 mainly spread from person to person through droplet transmission. Droplets are small pieces of saliva, which are produced when a person coughs or sneezes. Droplets cannot go through skin and can only lead to infection if they touch your mouth, nose or eye. Therefore, from a public health perspective it is very important to protect susceptible individuals from TB exposure by effectively reducing the contact rate between susceptible and infectious individuals.

There are so many ways that we can control COVID-19 transmission (i) wash your hands regularly with soap and water or rubbing an alcohol-based sanitizer into your hands because washing your hands kills viruses that may be on your hands, (ii) avoid touching your face as much as possible because virus containing droplets on your hands can be transferred to your eyes, mouth or nose where they can infect you, (iii) maintain at least 1.5 meters distance between yourself and anyone who is coughing or sneezing because if you are too close to someone you might breathe in droplets they cough or sneeze, (iv) make sure you and people around you follow good respiratory hygiene. Respiratory hygiene is important because droplets spread virus. By following good respiratory hygiene you catch any droplets that might be produced, and this protects the people around you from viruses including COVID-19. (v) Must wear a mask if you are sick with symptoms that might be due to COVID-19 or looking after someone who may have COVID-19.

However, estimation of transmission rates from different setting must be done with caution, as the pattern of an epidemic, the standard of care and, as a result, number of cases are time and setting dependent. For instance, very few cases have been reported so far in Bangladesh [23]. In this country, health system is very poor which leads to the fewer number of reported cases. Therefore, data from other countries, in particular the number of cases by date of COVID-19 onset is necessary to better understand the variability in cases across settings.

Our model determined that from the explicit expression for infectious individuals I and the basic reproduction number R_0_, it is clear that they are depending on transmission rates β, progression rates ω, recovery rates γ, and the force of infection saturates rate α. From the sensitivity analysis it is also clear that the most important parameter is transmission rate β followed by recovery rate γ. Therefore, to control and eradicate COVID-19 infection, it is important to consider the following strategies: (i) the first and most important strategy is to minimize the contact rates β with infected individuals by decreasing the values of β; (ii) the second-most important strategy is to increase the recovery rate γ of infective individuals through treatment. Therefore, we suggest the most feasible and optimal strategy to eliminate COVID-19 in six different countries including Australia, Italy, Spain, USA, UK and Canada are to reduce contact rates as well as increase the treatment rate that will be most effective way to reduce COVID-19 cases in the six countries. Finally, the application of proposed model and its related outputs can be extended into many other countries which are dealing with such a critical outbreak of COVID-19 to control this global pandemic disease.

## Data Availability

All data will be available upon request.

https://doi.org/10.6084/m9.figshare.12173589

## Ethical approval

This study based on aggregated COVID-19 surveillance data in Australia, Italy, Spain, USA, UK and Canada taken from the worldometer. No confidential information included because analyses were performed at the aggregate level. Therefore, no ethical approval is required.

## Data availability statement

All data will be available upon request.

## Contributors

AR conceived, designed and supervised the study. MAK contributed to the literature search and data collection. AR and MAK contributed to data analysis, table and figures, interpretation of results and writing of the manuscript.

## Acknowledgments

The authors would like to thank the editorial office for their quick response and valuable comments to produce the updated version.

